# Overdose mortality rates for opioids or stimulants are higher in males than females, controlling for rates of drug misuse: State-level data

**DOI:** 10.1101/2023.01.20.23284833

**Authors:** Eduardo R. Butelman, Yuefeng Huang, David H. Epstein, Yavin Shaham, Rita Z. Goldstein, Nora D. Volkow, Nelly Alia-Klein

**Author notes:** **Corresponding Author:** E.R. Butelman Ph.D., NARC Program, Department of Psychiatry, Icahn School of Medicine at Mount Sinai, One Gustave L. Levy Place, Box 1217, New York, NY 10029-6574.

## Abstract

**Importance:** Drug overdoses from opioids like fentanyl and heroin and stimulant drugs such as methamphetamine and cocaine are a major cause of mortality in the United States, with potential sex differences across the lifespan.

**Objective:** To determine overdose mortality for specific drug categories across the lifespan of males and females, using a nationally representative state-level sample.

**Design:** State-level analyses of nationally representative epidemiological data on overdose mortality for specific drug categories, across 10-year age bins (age range: 15-74).

**Setting:** Population-based study of Multiple Cause of Death 2020-2021 data from the Centers of Disease Control and Prevention (CDC WONDER platform).

**Participants:** Decedents in the United States in 2020-2021

**Main outcome measures:** The main outcome measure was sex-specific rates of overdose death (per 100,000) for: synthetic opioids excluding methadone (ICD-10 code: T40.4; predominantly fentanyl), heroin (T40.1), psychostimulants with potential for misuse, excluding cocaine (T43.6, predominantly methamphetamine; labeled “psychostimulants” hereafter), and cocaine (T40.5). Multiple regression analyses were used to control for ethnic-cultural background, household net worth, and sex-specific rate of misuse of the relevant substances (from the National Survey on Drug Use and Health, 2018-2019).

**Results:** For each of the drug categories assessed, males had greater overall overdose mortality than females, after controlling for rates of drug misuse. The mean male/female sex ratio of mortality rate for the separate drug categories was relatively stable across jurisdictions: synthetic opioids (2.5 [95%CI, 2.4-2.7]), heroin, (2.9 [95%CI, 2.7-3.1], psychostimulants (2.4 [95%CI, 2.3-2.5]), and cocaine (2.8 [95%CI, 2.6-2.9]). With data stratified in 10-year age bins, the sex difference generally survived adjustment for state-level ethnic-cultural and economic variables, and for sex-specific misuse of each drug type (especially for bins in the 25-64 age range). For synthetic opioids, the sex difference survived adjustment across the lifespan (i.e., 10-year age bins ranging from 15-74), including adolescence, adulthood and late adulthood.

**Conclusions and Relevance:** The robustly greater overdose mortality in males versus females for synthetic opioids (predominantly fentanyl), heroin, and stimulant drugs including methamphetamine and cocaine indicate that males who misuse these drugs are significantly more vulnerable to overdose deaths. These results call for research into diverse biological, behavioral, and social factors that underlie sex differences in human vulnerability to drug overdose.

**Key Points:** *Question:* What are the current national trends in overdose mortality from opioids (synthetic opioids such as fentanyl, and heroin) and stimulant drugs (psychostimulants such as methamphetamine and cocaine) for males and females, over the lifespan (overall range 15-74 years)?

*Findings:* State-level analyses of data from CDC for 2020-2021 indicate that after controlling for rates of drug misuse, males had significantly greater (2-3 fold) overdose mortality rates than females for synthetic opioids, heroin, psychostimulants and cocaine. These findings were generally consistent across the lifespan, studied as 10-year age bins (especially in the 25-64 age range).

*Meaning:* These data indicate that males who misuse opioids and stimulant drugs are considerably more vulnerable to overdose mortality, compared to females. This finding calls for research on the underlying biological, behavioral, and social factors.

## Introduction

Overdose deaths due to opioids (such as fentanyl and heroin), or stimulant drugs (such as methamphetamine and cocaine) have increased in recent years in the United States.^1-3^ This increase has accelerated since the onset of the COVID-19 pandemic (March 2020 and thereafter) for fentanyl and its analogs and stimulants such as methamphetamine, but not for heroin, prescription opioids, or methadone.^4-7^ This was due in part to increases in the distribution of synthetic opioids such as fentanyl, and their use to lace other drugs, including illicitly manufactured preparations disguised as prescription opioid analgesics, stimulant medications or benzodiazepines.^6, 8, 9^

Prevention of overdose death requires the identification of vulnerability factors, which are thought to include sex and age.^6^ Regarding sex as a biological variable, results from studies using rodent models are inconclusive due to conflicting results: some such studies have shown that female rats are more prone to escalate their intake of opioids or stimulant drugs,^10^ but a large number of other studies have not.^11^ Similarly, some studies in human cohorts have shown that females escalated their intake of opioids or stimulant drugs more rapidly than males,^12-15^ but several other studies have not.^16-18^ A recent comprehensive review did not find robust sex differences in vulnerability to craving and relapse.^11^ Nonetheless, as reviewed below, epidemiological data have suggested a sex difference in risk of overdose death, and those findings need to be followed up on.

Regarding age, epidemiological studies have suggested typical trajectories in which misuse of opioids or stimulant drugs begins in late adolescence or young adulthood and escalates thereafter.^18, 19^ However, the onset of misuse may also occur later in the lifespan, sometimes triggered iatrogenically.^20, 21^ Relatively recent data also point to an increase in older adults (aged 55 and older) seeking first-time treatment for OUD.^22^

Epidemiological studies show that males, compared to females, tend to have greater overdose mortality. For example, a recent report for 2019-2020, for 25 states and the District of Columbia, showed an increase in overdose mortality for all drugs combined, with a greater rate in males versus females.^23^ Similarly, a US nation-level analysis of mortality data showed an increase in overdoses from synthetic opioids (thus including fentanyl but not heroin) and psychostimulants (including methamphetamine but not cocaine) from March 2018 to March 2021, with highest mortality rates in males (especially from minoritized groups, e.g., African-American).^4^ A nation-level report on CDC data from 2017-2018 showed that males compared to females had greater overdose mortality for prescription opioids specifically, and for all opioids combined.^6^

In the analyses reported here, we followed up on those findings in greater depth, focusing on the sex difference across the lifespan, and asking whether its magnitude is consistent across US jurisdictions (50 states and district of Columbia), which can differ in major demographic and socioeconomic variables, levels of misuse, and illicit drug market factors.^24^ We examined overdose mortality separately for specific categories of opioid and stimulant drugs, due to their differing mechanisms of toxicity.^25^ Acute overdose mortality from opioids (mu-opioid receptor agonists) is primarily attributed to respiratory depression mediated by brainstem nuclei.^26, 27^ Overdose mortality from fentanyl and its analogs may be greater than that for heroin, due to greater potency, faster onset of action driven by high lipophilicity, and higher likelihood to trigger chest rigidity.^28-30^ It is not clear whether there are generalizable sex differences in sensitivity to respiratory depressant effects of mu-opioid agonists in humans.^31-34^ Epidemiological data do not directly address this question, but could provide evidence against it (or in favor of additional explanations) if the sex difference in overdose mortality is not consistent across state-level jurisdictions.

Acute overdose mortality by methamphetamine and cocaine in humans is primarily attributed to cardiovascular and neurological events such as malignant arrhythmias, strokes, or seizures.^35, 36^ Some, but not all, human studies reported that females had either more prolonged or more pronounced cardiovascular effects than males after administration of stimulant drugs.^37-40^ Again, epidemiological data cannot directly address questions of biological vulnerability, but could constrain hypotheses, depending on whether the sex difference in overdose mortality is consistent across states that differ in major demographic, socioeconomic and drug use factors.

In addition to sex differences in direct pathophysiological effects of the drug, factors that could account for stable variations in rates of overdose include: family and social interactions and related trauma,^41, 42^ propensity toward risky behaviors (e.g., injecting alone, taking large doses, or using untrusted suppliers),^43, 44^ and propensity to seek treatment.^45-49^ We expect that differences in these gendered behavioral factors would be largely consistent across drug categories and states in the US. Other contributing factors which could differ by state within the US include ethnic-cultural background, socioeconomic variables, illicit drug supply, and availability of evidence-based care.^8, 50-53^ Using state-level nationally representative CDC data for 2020-2021, the goal of this study was therefore to determine the extent to which sex differences in overdose mortality vary in four mutually exclusive drug categories (synthetic opioids, heroin, psychostimulants such as methamphetamine, and cocaine) and state, accounting for age. Different patterns of sex variability in overdose mortality would suggest (without proving or ruling out) different emphases on possible levels of causation.

## Methods

This was a study of de-identified publicly available data from the CDC “multiple cause mortality” data, obtained from the CDC WONDER platform for the years 2020-2021 (https://wonder.cdc.gov/mcd.html), which provides information based on death certificates in the United States. Overdose mortality data that were “suppressed” (due to n<10 per cell) or deemed “unreliable” in CDC WONDER were considered missing and were excluded from analysis. Data were analyzed in October 2022-February 2023. This study followed reporting guidelines from: Strengthening the Reporting of Observational Studies in Epidemiology (STROBE).

### Data set

The main analyzed outcome variable was crude death rate per 100,000 population for Drug Poisonings (ICD-10 codes for overdoses; including unintentional X40-X44, suicide X60-X64, homicide X85, and undetermined intents Y10-Y14). Data were obtained for four mutually exclusive categories: synthetic opioids other than methadone (“synthetic opioids” hereafter; this predominantly reflects fentanyl and its analogs, but also compounds from other chemical scaffolds; T40.4),^54^ heroin (T40.1), psychostimulants with potential for misuse (which predominantly reflects methamphetamine; T43.6) and for cocaine (T40.5). Data were stratified by sex and by six consecutive 10-year age bins (i.e., 15-24, 25-34, 35-44, 45-54, 55-64 and 65-74) obtained for 51 jurisdictions (50 states and the District of Columbia) for 2020-2021.^55^

### Nomenclature

This manuscript will use the terms “male” and “female” for data analyses ^56^, as this is the terminology used in death certificates, the root document for mortality data in CDC WONDER. For clarity, we use the term *“stimulant drugs”* to encompass compounds such as methamphetamine and cocaine overall, and use the term “psychostimulants” for the CDC overdose category (T43.6) that predominantly reflects methamphetamine. As stated above, cocaine overdoses (T40.5) are analyzed in a separate category from psychostimulants.

### Data Analyses

All analyses were carried out with GraphPad Prism V.9, using jurisdiction-level data. Univariate analyses included Spearman correlations on overdose mortality data (crude death rate per 100,000 population) for males and females, per jurisdiction. Mortality and misuse ratios in males/females were compared with Wilcoxon tests. Univariate mixed-effects ANOVAs (sex x age bin) then analyzed log-transformed overdose mortality rate across the lifespan, stratified in six consecutive 10-year age bins; significant effects were followed with post-hoc Bonferroni tests.

Separately for each drug category (all ages combined in the range 15-74 years), multivariable analyses were then carried out with multiple linear (least squares) regressions, to assess the effect of sex and selected potential covariates on the outcome, log-transformed death rates. Thus, the independent variables included: sex (M/F), percent of the population who is white, percent of the population who is black (from the 5-year American Community Survey for 2020 from the U.S. Census Bureau; https://worldpopulationreview.com/states/states-by-race), and median household net worth in 2020 (www.census.gov/data/tables/2020/demo/wealth/state-wealth-asset-ownership.html). Using data from the National Survey on Drug Use and Health (NSDUH) for 2018-2019, we also controlled for overall levels of misuse of the relevant drugs (https://rdas.samhsa.gov/#/survey/NSDUH-2018-2019-RD02YR). Thus, the multiple linear regressions for overdose mortality by synthetic opioids and heroin controlled for the rate of past-year misuse of opioids, by sex (item “OPINMYR” in the NSDUH survey). Likewise, the regressions for overdose mortality caused by psychostimulants or cocaine controlled for the rate of past-year misuse of these drugs, by sex (items “AMMEPDPYMU” and “COCMYR” in the NSDUH survey, respectively). Using the NSDUH survey, we were not able to perform state- and sex-specific analyses of relevant substance use disorders (as opposed to misuse), because of frequent data suppression after stratification. The overdose mortality data were also stratified by 10-year age bins (overall range: 15-74 years), and multiple regressions were run separately for each age bin.

In the ANOVAs and multiple regressions, outliers for the outcome variable (greater than ±1.96 standard deviations from the mean were removed, using Grubbs test at the 5% alpha level of significance. Also, if there were <10 jurisdictions in which overdose mortality data were not available for both males and females for an age bin and drug category, the data set was excluded from analysis, in order to maintain representativeness. The overall alpha level of significance was set at the p=0.05 level, two-tailed.

In a supplementary analysis, we also examined analogously overdoses caused by both synthetic opioids *and* psychostimulants (a common poly-drug pattern of overdose) (Supplement).^3, 57^

## Results

### Overall overdose mortality profiles (all ages combined)

With all ages combined (overall range 15-74 years), there was a broad range of overdose mortality rates for the four drug categories, across jurisdictions (Figures 1 and 2; lower panels, and eTable 1). There were positive Spearman correlations, nearing unity, in the rate of overdose mortality for males and females in the same jurisdiction, for each of the drug categories (Spearman correlations ranged from +0.94 to +0.97; all p-values were <0.0001) (see lower panels of Figure 1 for synthetic opioids and heroin, and Figure 2 for psychostimulants and cocaine). Simple linear regressions provided a strong fit for these data (R^2^ values of 0.95, 0.88, 0.92 and 0.95, for synthetic opioids, heroin, psychostimulants and cocaine respectively). The mean ratios for mortality in males/females (all ages combined) were in the 2.4-2.9 range: ratios were 2.5 (95%CI: 2.4-2.7) for synthetic opioids, 2.9 (95%CI: 2.7-3.1) for heroin, 2.4 (95%CI: 2.3-2.5) for psychostimulants and 2.8 (95%CI: 2.6-2.9) for cocaine.

**Figure 1:**
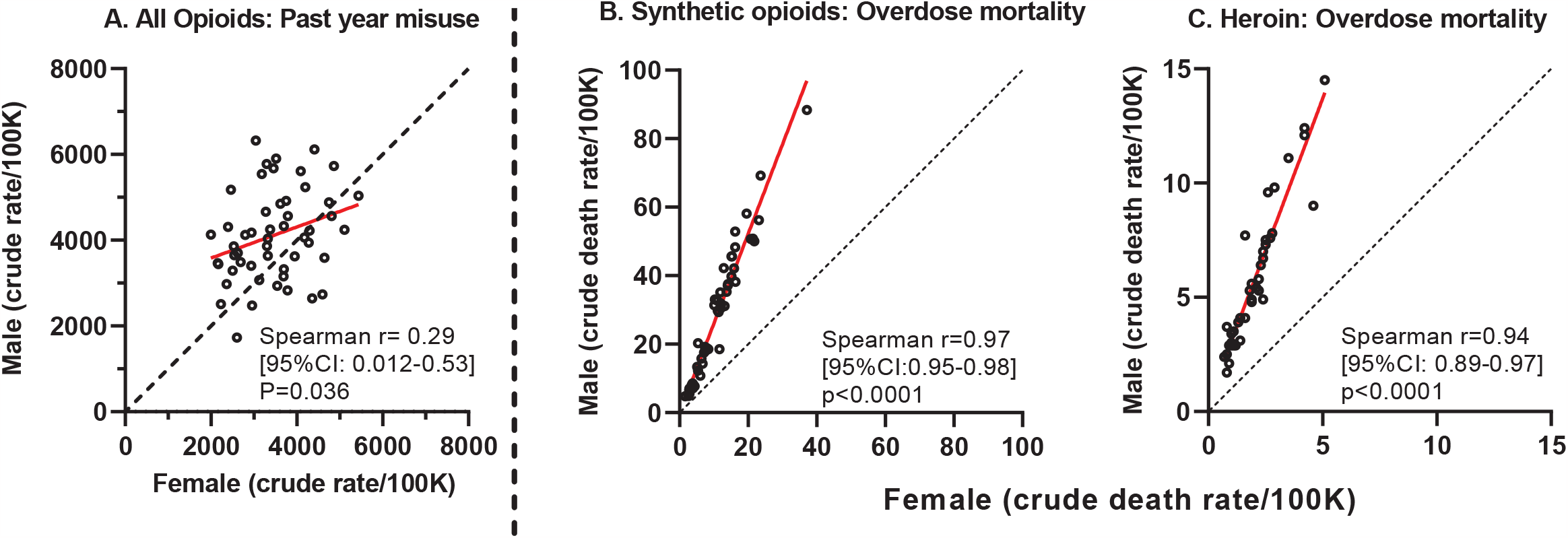
Opioid misuse and overdose mortality. Upper panel: Past-year misuse of any opioid (upper panel) in males and females, all ages combined. Lower panels: Overdose mortality due to synthetic opioids and heroin in males and females; all ages combined. For all panels, each symbol (open circles) is one jurisdiction. Note different axis ranges in the panels. Spearman correlations for males and females in each jurisdiction are included. The red line indicates the best-fit simple linear regression (not shown in the upper panel, as the 95%CI of the slope overlapped with 0). The dashed black line indicates identity.

**Figure 2:**
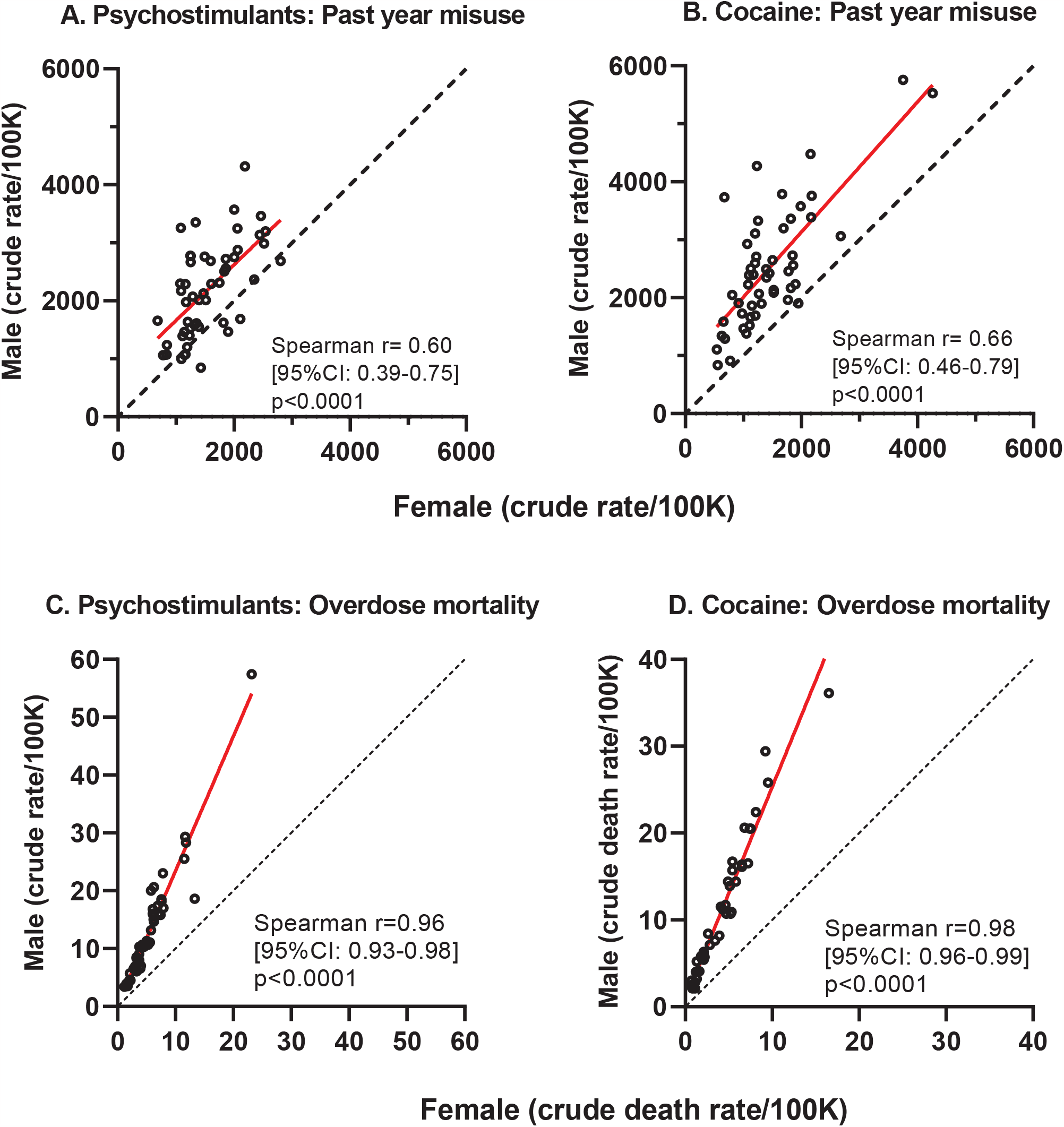
Psychostimulant and cocaine misuse and overdose mortality. Upper panels: Past-year misuse of psychostimulants and cocaine in males and females, all ages combined. Lower panels: Overdose, mortality due to synthetic opioids and heroin in males and females; all ages combined. For all panels, each symbol (open circles) is one jurisdiction. Note different axis ranges in the panels. Spearman correlations for males and females in each jurisdiction are included. The red line indicates the best-fit simple linear regression. The dashed black line indicates identity.

In contrast, the correlations for past-year drug misuse between males and females within jurisdictions were of smaller magnitude, ranging from Spearman r value of 0.29 for opioids, to 0.60 for psychostimulants, and 0.66 for cocaine) (Figures 1 and 2; upper panels). Also, the male/female ratios of misuse were significantly smaller than the male/female ratios for overdose mortality, for each of these drug categories (see Figure 3).

**Figure 3:**
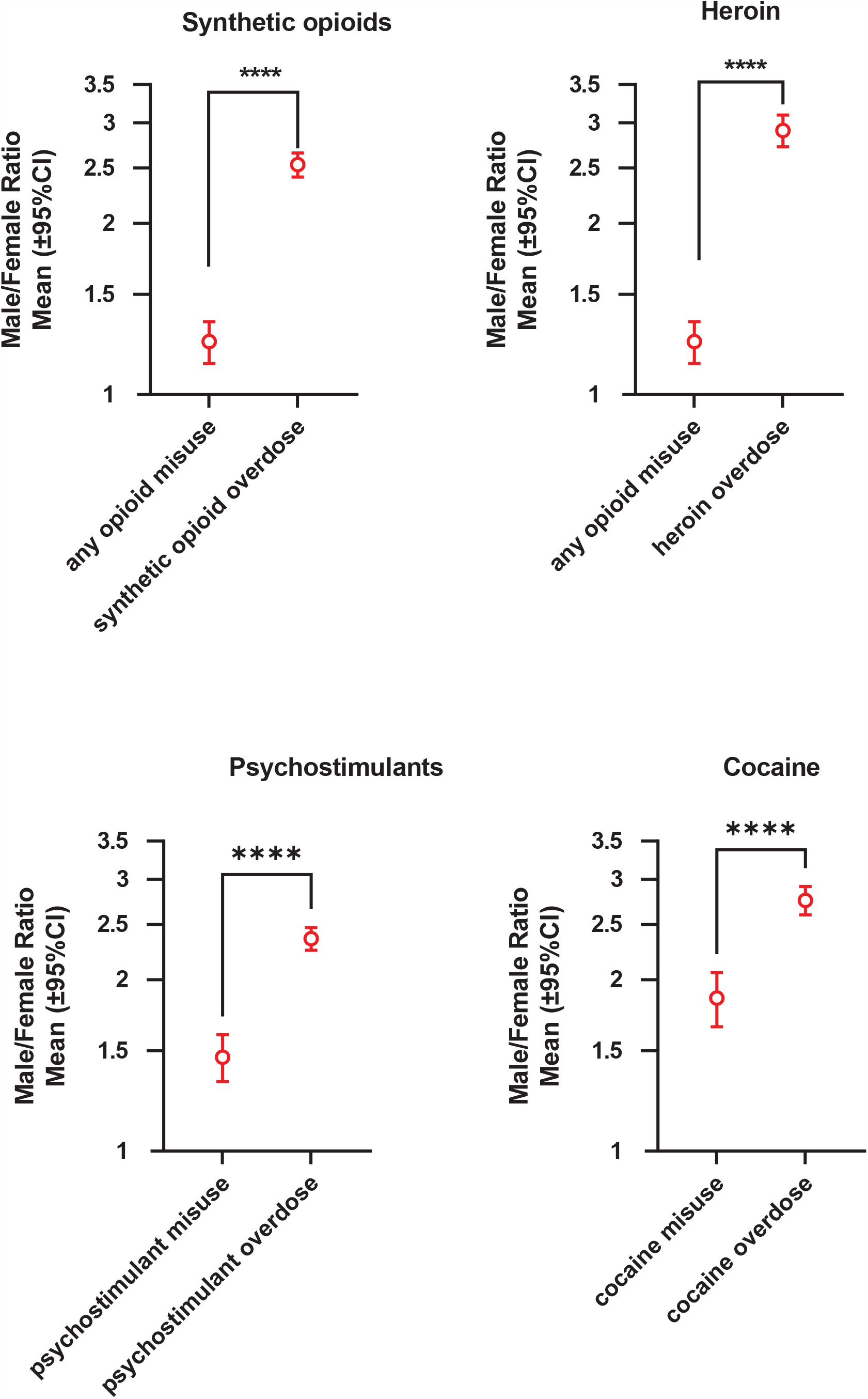
Male/Female ratios for misuse versus overdose mortality. Male/Female ratios for past-year misuse of relevant substances and for overdose mortality (shown in logarithmic axis). Data were analyzed with Wilcoxon’s tests, followed by Bonferroni analysis where appropriate (i.e., based on opioid misuse data being used both in synthetic opioid and heroin panels). P-values are shown as **** (p=0.0001) and *** (p=0.001). Jurisdictions with missing data were excluded from the analysis.

### Univariate analyses of overdose mortality across the lifespan

Separate 2×2 ANOVAs (sex x age bin) were carried out for each drug category. For synthetic opioids, psychostimulants and cocaine, there were significant main effects of sex and age bin, and also a sex by age bin interaction (see Figure 4 and eTables 2 and 3). For heroin, there were significant main effects of sex and age bin, but the interaction did not reach significance. Post-hoc Bonferroni tests for sex at each age bin showed that with few exceptions (typically for the youngest and oldest age bins, where the lowest rates were observed), males had greater mortality than females.

**Figure 4:**
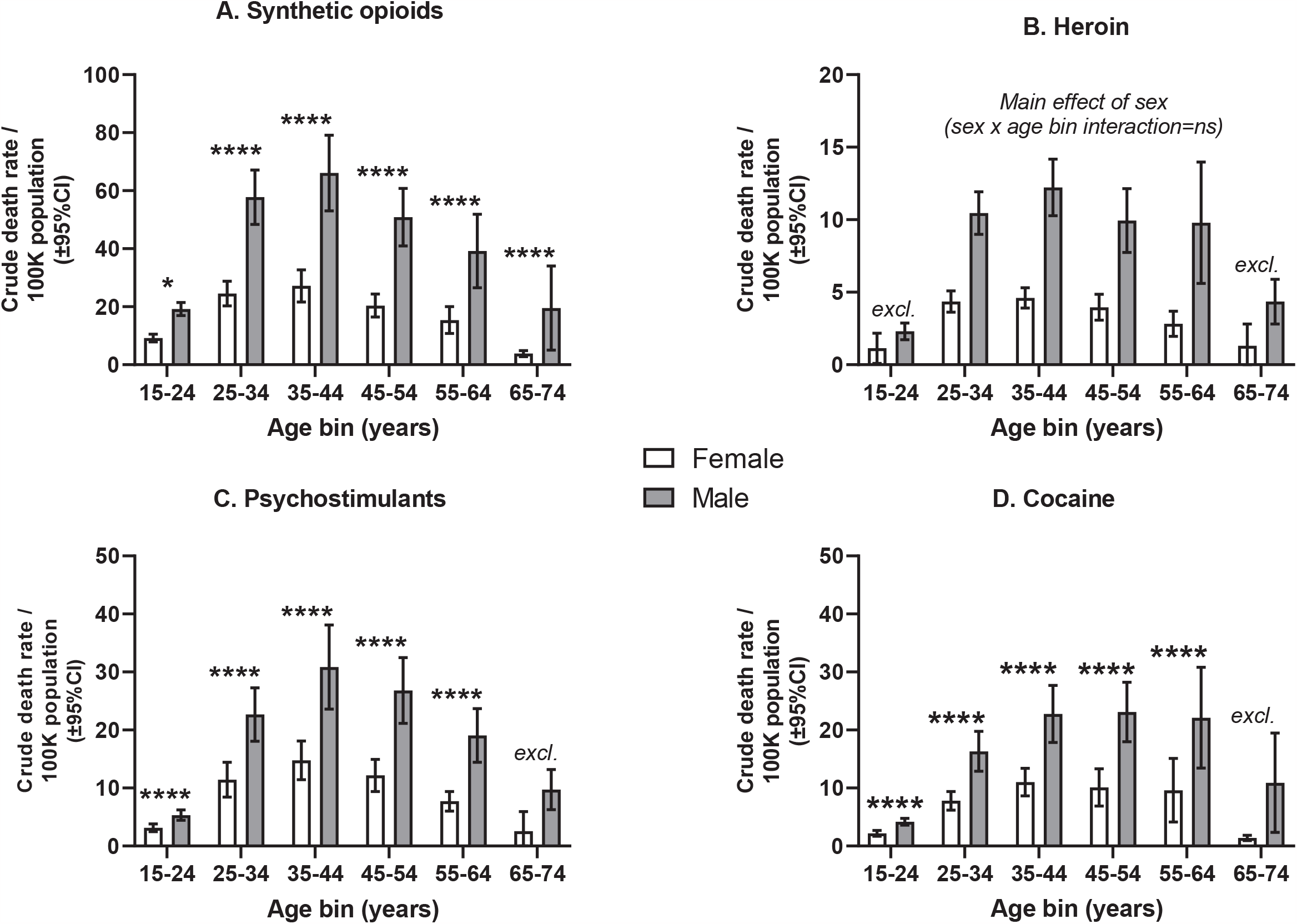
Summary of overdose death rates for different drug categories across the lifespan. Overdose mortality in males and females, for consecutive 10-year age bins. Note the different Y-axis ranges in the different panels. Means and 95%CI are calculated across 51 jurisdictions. Log –transformed overdose data were analyzed with univariate mixed-effects effects ANOVAs (sex x age bin). Bonferroni post-hoc tests for sex at each age bin are shown with black brackets (* is p<0.05; **** is p<0.0001). The label “excl.” indicates that the age bin was excluded from ANOVA analysis due to insufficient available data (see Methods and eTable 2). Full ANOVA table is present in the Supplement (eTable 3)

### Multivariable analyses

Multiple linear regressions were conducted for each drug category separately, with all ages combined (age range 15-74). Sex remained a significant factor for overdose mortality for all drug categories, after adjustment for the jurisdiction-level sex-specific rate of misuse of the relevant drugs, and for demographic variables (distribution of major ethnic-cultural groups and median household net worth) (see eTable 4, and Figure 5).

**Figure 5:**
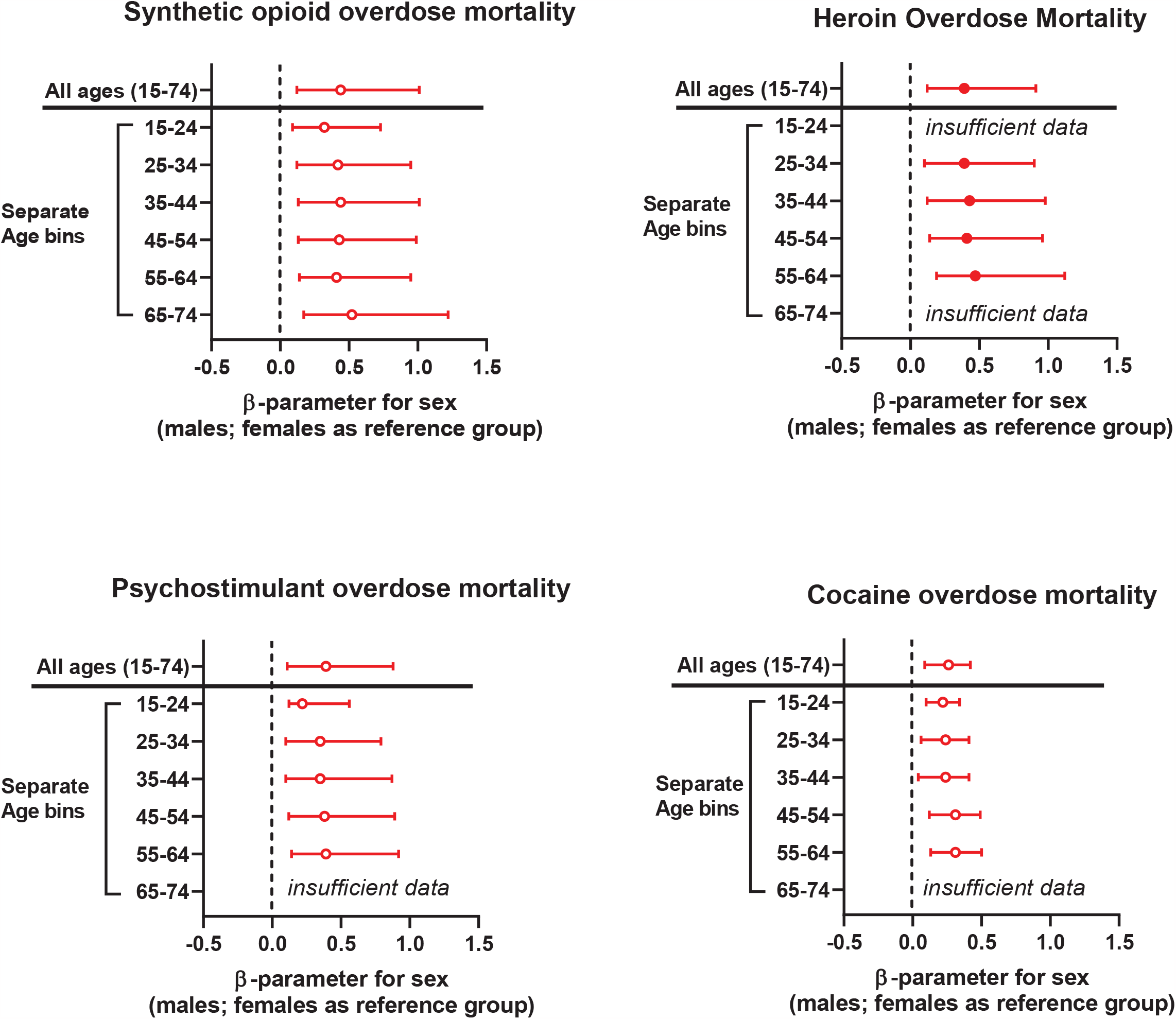
Sex as a factor in overdose mortality after adjustment for levels of drug misuse and demographic variables. β(beta)-parameter for sex as a factor in overdose mortality, from multiple linear regressions. The outcome was log-transformed overdose mortality per 100,000 population. The data were adjusted for jurisdiction-specific levels of misuse in males and females, and for three demographic variables (%White, %African American, and household assets). Full regression data are presented in the Supplement (eTable 4, eTable 5).

Similar multiple linear regressions were then conducted, after stratification into 10-year age bins. For all drug categories and age bins, sex remained a significant factor in overdose mortality rates after adjustment. Of note, that included the 55-64 age bin (see eTable 5 and Figure 5).

### Supplementary analysis of overdose mortality caused by *both* synthetic opioids *and* psychostimulants

In a follow-up analysis, we examined overdose mortality caused by a commonly reported pattern of polydrug use: combining synthetic opioids and psychostimulants (i.e., ICD-10 codes T40.4 and T43.6 reported together as multiple causes of mortality).^3, 57, 58^ This outcome generally followed a similar profile across the lifespan, with males having greater mortality rates than females (eFigure 1, eTable 6 and eTable 7).

## Discussion

This was a nationally representative state- and age-specific examination of differences between males and females in overdose mortality due to opioid and stimulant drugs, for 2020-2021. The study showed that for these drug categories, across the lifespan, after controlling for sex-specific levels of past-year drug misuse and for major demographic factors, males died from overdoses at an approximately 2-3 greater rate than females. Studies from several other industrialized countries do show greater drug-induced mortality for males versus females ^59, 60^. However, to the best of our knowledge, recent comparative data for these different drugs, stratified by age, are not published for a broad set of countries.

For synthetic opioids, the effect of sex on overdose mortality survived adjustment in all 10-year age bins (overall range 15-74 years). For the other drug categories under study (heroin, psychostimulants and cocaine), this effect of sex survived adjustment in age bins in the 25-64 age range. Therefore, for all the drugs studied here, greater overdose mortality rates were observed in males versus females in the 55-64 age bin, considered to be in the post-menopausal range for the population. ^61^

The robustness of the overall sex difference in overdose mortality, even after controlling for different propensity to misuse, and across state-level jurisdictions, shows that males in the US are reliably at greater risk of fatal overdose than females. Mechanistic explanations cannot be directly derived from these epidemiological data, but some processes can be accorded greater likelihood of accounting for most of the sex difference. Sex-specific biological vulnerability to the direct toxic effects of the drugs, for example, cannot be ruled out. However, this sex-specific vulnerability would have to be shared across mu-opioid agonists and stimulant drugs, whose toxic effects are highly distinct from each other, in terms of pharmacodynamic targets and pathophysiological mechanisms. Even within these major drug categories, the pharmacodynamics and pathophysiological effects of specific mu-opioid agonists such as fentanyl versus morphine derivatives such as heroin,^29, 30^ and methamphetamine versus cocaine,^62, 63^ are not identical.

There are sex differences in the propensity to development of substance-use disorders (SUDs), but they appear unlikely to account for the observed difference in mortality rates. Nation-level data from the 2021 NSDUH^64^ show that the overall male/female ratio in prevalence of any past-year SUD (all drugs combined) was 1.3 (9.8% of males versus 7.4% of females)—a considerably smaller sex difference than the one we observed in mortality rates. We noted earlier than other, more culture-bound differences between men and women could include differential propensity toward risky behaviors, above and beyond any risk inherent in misusing an opioid or stimulant—e.g., injecting alone,^43, 44^ taking large doses, or using untrusted suppliers.

To explore the possibility that such a broader difference in risky behaviors underlies mortality, we undertook a follow-up analysis of the same CDC WONDER dataset for 2020-2021, for the broad category of “Transport Accidents” (ICD-10 codes: V01-V99, thus including decedent drivers, passengers and pedestrians). Using state-level data, the male/female ratio in fatalities due to transport accidents is similar in magnitude and consistency to the ratios observed above for overdose mortality (i.e., the mean male/female ratio for transport accidents was 2.68; 95%CI: 2.60-2.76, with a correlation for males and females in the same jurisdiction nearing unity; Spearman r=0.97; p<0.0001). One caveat is that these CDC data on transport accidents combine instances in which the decedent was or was not determined to be responsible for the event. A published study, however, suggests that men are 2-3 fold more likely than women to be responsible for motor collisions, and are not more likely to be passive victims of motor collisions.^65^ This is consistent with recent nationally representative US data sets (e.g., from 2020) showing that male drivers who died as a result of traffic accidents had a greater likelihood of engaging in risky behaviors such as speeding, compared to female drivers ^66^. We suspect that this type of sex difference is a valuable starting point for the exploration of both a major mechanism and a modifiable risk factor in overdose deaths, but this explanation does not exclude important roles for other biological, behavioral or social factors.

### Limitations

We focused on drug categories that greatly contribute to overdose deaths in the US at this time^3, 4, 67^, but there is a need for similar examination of sex differences in overdose death from prescription opioids, benzodiazepines and alcohol.^6, 68^ Also, we cannot fully exclude the possibility that some of the sex differences in overdose mortality or misuse are due to reporting biases.^69, 70^ In addition, state- and sex-level data on drug misuse from NSDUH for years after 2019 are not are not currently publicly available. Also, it should be noted that the opioid misuse data in NSDUH (item OPINMYR) can encompass other compounds, in addition to synthetic opioids and heroin.

## Conclusions

We performed a state-level analysis of recent nationally representative data, and found that males, compared to females, have a significantly greater rate of overdose mortality from synthetic opioids, heroin, and stimulant drugs such as methamphetamine and cocaine. An important finding in our study is that greater mortality in males was evident even after controlling for levels of misuse. These sex differences were observed across the lifespan (e.g., in the range of 25-64 years of age), and were stable across states that have broad range of environmental variables. Overall, these findings underscore that sex differences are important targets for investigation at multiple biological and behavioral levels, potentially leading to optimized prevention and intervention approaches to mitigate the risk of overdose mortality, at different stages across the lifespan.

## Data Availability

All data produced in the present study are available upon reasonable request to the authors.

https://rdas.samhsa.gov/#/survey/NSDUH-2018-2019-RD02YR

https://www.census.gov/data/tables/2020/demo/wealth/state-wealth-asset-ownership.html

https://worldpopulationreview.com/states/states-by-race

## Abbreviations

95%CI: 95% confidence interval
CDC: Centers for Disease Control and Prevention
ICD-10: International Classification of Diseases, 10^th^ Edition
Psychostimulants: Abbreviation for ICD-10 category T43.6; “psychostimulants with potential for misuse, excluding cocaine”
SUD: Substance use disorders

## Author Contributions

All authors contributed to the writing and editing process, including text and data analysis quality, and approved submission of the manuscript. All authors agree to be accountable for all aspects of the manuscript, including accuracy and integrity, and investigation and resolution of any potential discrepancies.

## Data Availability Statement

This study used publicly available de-identified information, data sources are described in the Methods section.

## Funding

This work was supported by NIDA U01DA053625 (ERB), NIDA 1RO1DA048301-01A1 (RZG) and NIDA 1RO1DA049547 (NAK), and by Intramural Research Program funds of NIDA (YS and DHE).

## Competing Interests

The authors have nothing to disclose.

## Notes

### Competing Interest Statement

The authors have declared no competing interest.

### Funding Statement

NIH-NIDA U01DA053625 (ERB), and NIDA 1RO1DA048301-01A1 (RZG), NIDA 1RO1DA049547 (NAK)

### Author Declarations

Source data were openly available at the beginning of the study, as de-identified data from the U.S. CDC multiple cause mortality data, obtained from the CDC WONDER platform (https://wonder.cdc.gov/mcd.html)

### Summary of Updates

Text of Introduction and Discussion has been edited. Figures have been updated and data analyses have been updated.

