## Supplemental Data for "Overdose mortality rates for opioids or stimulants are higher in males than females, controlling for rates of drug misuse: State-level data"

**Supplement**

**eTable 1: Overdose mortality rates for all ages combined (age range 15-74)**

**(total 51 jurisdictions; 50 states and District of Columbia)**

|  | **Synthetic opioids** | | **Heroin** | | **Psychostimulants** | | **Cocaine** | |
| --- | --- | --- | --- | --- | --- | --- | --- | --- |
| **Sex** | **Male** | **Female** | **Male** | **Female** | **Male** | **Female** | **Male** | **Female** |
| **Mean^a^ (±95%CI)** | 29.0  (23.7-34.3) | 11.1  (9.2-13.1) | 5.5  (4.6-6.5) | 2.0  (1.7-2.4) | 13.0  (10.4-15.6) | 5.6  (4.5-6.7) | 10.6  (8.2-13.0) | 4.2  (3.2-5.2) |
| **N** | 51 | 51 | 45 | 39 | 50 | 48 | 45 | 42 |
| **Missing^b^** | 0 | 0 | 6 | 12 | 1 | 3 | 6 | 9 |

^a^Overdose mortality rate per 100,000 population, unadjusted

^b^Missing values occurred when data for a jurisdiction were “suppressed” or considered “unreliable” in CDC WONDER

**eTable 2: Overdose mortality rates per 10-year age bin (age range 15-74)**

**(total 51 jurisdictions; 50 states and District of Columbia)**

| **Age Bin**  **(years)** |  | **Synthetic opioids** | | **Heroin** | | **Psychostimulants** | | **Cocaine** | |
| --- | --- | --- | --- | --- | --- | --- | --- | --- | --- |
|  | **Sex** | **Male** | **Female** | **Male** | **Female** | **Male** | **Female** | **Male** | **Female** |
| **15-24** | **Mean^a^ (±95%CI)** | 19.2  (17.0-21.5) | 9.2  (7.9-10.6) | 2.3  (1.7-2.9) | 1.1  (0.1-2.2) | 5.3  (4.4-6.2) | 3.2  (2.5-3.8) | 4.2  (3.6-4.8) | 2.2  (1.7-2.7) |
|  | **N^b^** | 43 | 33 | 9 | 3 | 31 | 20 | 23 | 6 |
|  | **Missing^c^** | 8 | 18 | 42 | 48 | 20 | 31 | 28 | 45 |
| **25-34** | **Mean^a^ (±95%CI)** | 57.8  (48.4-67.1) | 24.5  (20.3-28.8) | 10.5  (9.0-11.9) | 4.4  (3.6-5.1) | 22.7  (18.0-27.3) | 11.4  (8.4-14.4) | 16.3  (12.9-19.8) | 7.8  (6.2-9.4) |
|  | **N** | 48 | 44 | 37 | 25 | 44 | 37 | 38 | 29 |
|  | **Missing^b^** | 3 | 7 | 14 | 26 | 7 | 14 | 13 | 22 |
| **35-44** | **Mean^a^ (±95%CI)** | 66.1  (53.0-79.2) | 27.2  (21.7-32.7) | 12.2  (10.3-14.2) | 4.6  (3.9-5.3) | 30.9  (23.6-38.1) | 14.8  (11.5-18.1) | 22.8  (17.8-27.7) | 11.0  (8.6-13.4) |
|  | **N** | 47 | 43 | 37 | 22 | 47 | 38 | 39 | 29 |
|  | **Missing^b^** | 4 | 8 | 14 | 29 | 4 | 13 | 12 | 22 |
| **45-54** | **Mean^a^ (±95%CI)** | 50.9  (41.0-60.8) | 20.4  (16.4-24.8) | 9.4  (7.5-12.1) | 5.0  (3.1-4.9) | 36.8  (21.2-32.5) | 12.2  (9.4-20.0) | 23.1  (18.0-28.2) | 10.1  (6.9-13.3) |
|  | **N** | 44 | 43 | 34 | 21 | 42 | 37 | 38 | 29 |
|  | **Missing^b^** | 7 | 8 | 17 | 30 | 9 | 14 | 13 | 22 |
| **55-64** | **Mean^a^ (±95%CI)** | 39.2  (26.6-51.9 | 15.4  (10.8-20.0) | 9.8  (5.6-14.0) | 2.8  (2.0-3.7) | 19.1  (14.5-23.7) | 7.7  (6.0-9.4) | 22.1  (13.4-30.8) | 9.6  (4.1-15.1) |
|  | **N** | 43 | 40 | 30 | 13 | 42 | 33 | 37 | 24 |
|  | **Missing^b^** | 8 | 11 | 21 | 38 | 9 | 18 | 14 | 27 |
| **65-74** | **Mean^a^ (±95%CI)** | 19.5  (5.0-34.0) | 3.8  (2.8-4.9) | 4.4  (2.8-5.9) | 1.3  (0.6-2.0) | 9.7  (6.3-13.2) | 2.6  (1.1-4.1) | 10.9  (2.3-19.5) | 1.4  (0.9-1.9) |
|  | **N** | 29 | 12 | 11 | 3 | 20 | 3 | 23 | 6 |
|  | **Missing^b^** | 22 | 39 | 40 | 48 | 31 | 48 | 28 | 45 |

^a^Overdose mortality rate per 100,000 population, unadjusted

^b^Note: If N<10 for either males or females for an age bin in a drug category, the entire age bin was excluded from analyses (both ANOVA or multiple linear regressions) for that drug category.

^c^Missing values for an age bin occurred when data for a jurisdiction were “suppressed” or considered “unreliable” in CDC WONDER

**eTable 3:** Univariate mixed-effects ANOVAs (sex x age bin) of overdose mortality for specific drug categories.

| **Drug Category** | **Sex**  **Main effect** | **Age bin**  **Main effect** | **Interaction**  **(Sex x**  **Age bin)** | **Significant post-hoc tests**  **for sex x age bin** |
| --- | --- | --- | --- | --- |
| Synthetic opioids | F (1, 209) = 2291;  p<0.0001 | F (5, 248) = 31.9; p<0.0001 | F (5, 209) = 6.4;  p<0.0001 | Male>Female at age bins:   - 15-24 - 25-34 - 35-44 - 45-54 - 55-64 - 65-74 |
| Heroin  (excluded 15-24 and 65-74 age bins^a^) | F (1, 77) = 1209;  p<0.0001 | F (3, 134) = 4.9;  P=0.0031 | F (3, 77) = 2.1;  NS | - N/A   (sex x age bin  interaction not significant) |
| Psychostimulants  (excluded 65-74 age bin^a^) | F (1, 160) = 1736;  p<0.0001 | F (4, 201) = 30.4;  p<0.0001 | F (4, 160) = 16.4;  p<0.0001 | Male>Female at age bins:   - 15-24 - 25-34 - 35-45 - 45-54 - 55-64 |
| Cocaine  (excluded 65-74 age bin^a^) | F (1, 119) = 1295;  p<0.0001 | F (4, 170) = 17.6;  P<0.0001 | F (4, 119) = 5.3;  p=0.0006 | Male>Female at age bins:   - 15-24 - 25-34 - 35-45 - 45-54 - 55-64 |

^a^Age bins were excluded from analysis if there were <10 jurisdictions with data in both males and females.

**eTable 4:** Multiple linear regressions for log mortality rate for different drug categories: **all ages (age range 15-74)**

| **Significant β (beta) parameter Estimates [95%CI] and p-value** | | | | | | Model R^2^ (DF) |
| --- | --- | --- | --- | --- | --- | --- |
| Drug Category | Sex (Men);  Women as reference | State Median  Household  Net Worth ($) | State  %White | State  %Black | State and sex  past year opioid, stimulant or cocaine misuse, as applicable  (log rate)^a^ |  |
| Synthetic opioids | β**=**0.44  [0.32-0.57]  p<0.0001  eta^2^=0.30^b^ | NS | NS | β**=**0.010  [0.0040-0.17]  P=0.0016 | NS | 0.41  (94) |
| Heroin | β**=**0.39  [0.27-0.52]  p<0.0001  eta^2^=0.31 | NS | NS | NS | NS | 0.42  (76) |
| Psychostimulants | β**=**0.39  [0.28-0.49]  p<0.0001  eta^2^=0.31 | β=−1.93 X 10^-6^  [-2.69 X 10^-6^ –  -1.17^-6^]  p<0.0001 | β=−0.0068  [-0.0013 –  -0.0013]  p=0.016 | β=−0.014  [-022 –  -0.0070]  p=0.0002 | NS | 0.51  (90) |
| Cocaine | β**=**0.26  [0.087-0.42]  p=0.0034  eta^2^=0.11 | NS | β**=**0.012  [0.0038-0.020]  p=0.0043 | β**=**0.021  [0.012-0.031]  P<0.0001 | β**=**0.65  [0.22-1.07]  p=0.0035 | 0.45  (80) |

Outcome: Log-transformed mortality rate per 100K population

^a^State- and sex- level of past year opioid (for synthetic opioids and heroin), psychostimulant or cocaine misuse was from NSDUH (2018-2019), item “OPINMYR”, “AMMEPDPYMU” and “COCYR” respectively, log-transformed rate per 100,000 population.

**eTable 5:** Multiple linear regressions for log mortality rate, by 10-year age bin

| **eTable 5.1**  **Synthetic Opioids** | **Significant β (beta) parameter Estimates [95%CI] and p-value** | | | | | Model R^2^ (DF) |
| --- | --- | --- | --- | --- | --- | --- |
| Age bin | Sex (Men);  Women as reference | State Median  Household  Net Worth ($) | State  %White | State  %Black | State and sex  past year opioid misuse (log rate)^a^ |  |
| 15-24 | β**=**0.32  [0.23-0.41]  p<0.0001  eta^2^=0.39^b^ | NS | NS | NS | NS | 0.48  (69) |
| 25-34 | β**=**0.42  [0.30-0.53]  p<0.0001  eta^2^=0.31 | NS | β**=**0.011  [0.0031-0.018]  p=0.0067 | β**=**0.012  [0.0038-0.021]  p=0.0052 | NS | 0.43  (83) |
| 35-44 | β**=**0.44  [0.31-0.57]  p<0.0001  eta^2^=0.31 | NS | β**=**0.013  [0.0061-0.020]  p=0.0003 | β**=**0.017  [0.0082-0.025]  p=0.0002 | -0.73  [-1.31 - -0.15]  P=0.014 | 0.45  (81) |
| 45-54 | β**=**0.43  [0.30-0.56]  P<0.0001  eta^2^=0.30 | β**=**1.05 X 10^-6^  [1.49 X 10^-7^ –  1.94 X 10^-6^]  P=0.023 | β**=**0.014  [0.0056-0.023]  p=0.0015 | β**=**0.021  [0.011-0.030]  p=0.0001 | -0.75  [-1.35 - -0.16]  P=0.014 | 0.44  (79) |
| 55-64 | β**=**0.41  [0.27-0.54]  p<0.0001  eta^2^=0.28 | β**=**1.19 X 10^-6^  [1.97 X 10^-7^ –  2.81 X 10^-6^]  P=0.020 | NS | β**=**0.016  [0.0051-0.026]  p=0.0042 | -0.89  [-1.40 - -0.29]  P=0.0042 | 0.37  (76) |
| 65-74 | β**=**0.52  [0.35-0.70]  p<0.0001  eta^2^=0.47 | β**=**1.40 X 10^-6^  [7.50 X 10^-8^ –  2.72 X 10^-6^]  P=0.039 | NS | β**=**0.015  [0.0018-0.028]  p=0.028 | NS | 0.58  (34) |

Outcome: Log-transformed mortality rate per 100K population

^a^State level of past year opioid misuse was from NSDUH, item “OPINMYR” (2018-2019), log rate / 100,000 per sex.

^b^eta^2^ for sex (i.e., sum of squares_sex_ / sum of squares_total_); to measure effect size for the main variable of interest ^65^.

| **eTable 5.2**  **Heroin** | **Significant β (beta) parameter Estimates [95%CI] and p-value** | | | | | Model R^2^ (DF) |
| --- | --- | --- | --- | --- | --- | --- |
| Age bin | Sex (Men);  Women as reference | State Median  Household  Net Worth ($) | State  %White | State  %Black | State and sex  past year opioid misuse (log rate)^a^ |  |
| 15-24 | Insufficient data for regression^c^ | | | | | N/A |
| 25-34 | β**=**0.39  [0.29-0.51]  p<0.0001  eta^2^=0.46 | NS | NS | NS | NS | 0.51 (54) |
| 35-44 | β**=**0.43  [0.31-0.55]  p<0.0001  eta^2^=0.48 | NS | NS | NS | NS | 0.53  (51) |
| 45-54 | β**=**0.41  [0.27-0.55]  p<0.0001  eta^2^=0.40 | NS | NS | NS | NS | 0.47  (48) |
| 55-64 | β**=**0.47  [0.28-0.65]  p<0.0001  eta^2^=0.37 | NS | NS | NS | NS | 0.51 (34) |
| 65-74 | Insufficient data for regression^c^ | | | | | N/A |

^a^State level of past year opioid misuse was from NSDUH, item “OPINMYR” (2018-2019), log rate / 100,000 per sex.

^c^Less than 10 States had available overdose mortality data in both males and females (see Methods and eTable 2).

| **eTable 5.3**  **Psychostimulants** | **Significant β (beta) parameter Estimates [95%CI] and p-value** | | | | | Model R^2^ (DF) |
| --- | --- | --- | --- | --- | --- | --- |
| Age bin | Sex (Men);  Women as reference | State Median  Household  Net Worth | State  %White | State  %Black | State and sex  past year stimulant misuse  (log rate)^d^ |  |
| 15-24 | β**=**0.22  [0.097-0.34]  P=0.0003  eta^2^=0.20 | NS | NS | NS | NS | 0.33 (44) |
| 25-34 | β**=**0.35  [0.25-0.44]  p<0.0001  eta^2^=0.29 | β**=**-1.92 X 10^-6^  [-2.63 X 10^-6^ –  -1.20 X 10^-6^]  p=0.0001 | NS | NS | NS | 0.60 (73) |
| 35-44 | β**=**0.35  [0.25-0.52]  p<0.0001  eta^2^=0.34 | β**=**-1.64 X 10^-6^  [-7.33 X 10^-7^ –  -2.56 X 10^-6^]  p=0.0006 | NS | NS | NS | 0.47  (77) |
| 45-54 | β**=**0.38  [0.26-0.51]  p<0.0001  eta^2^=0.28 | β**=**-2.19 X 10^-6^  [-3.19 X 10^-6^ –  -1.19 X 10^-6^]  P<0.0001 | NS | β**=**-0.019  [-0.028 –  -0.0089]  p=0.0003 | NS | 0.48  (71) |
| 55-64 | β**=**0.39  [0.25-0.53]  P<0.0001  eta^2^=0.22 | β**=**-1.90 X 10^-6^  [-7.81 X 10^-7^ –  -3.02 X 10^-6^]  p=0.0012 | β**=**-0.011  [-0.019 –  -0.0035]  p=0.0053 | β**=**-0.028  [-0.039 –  -0.018]  P<0.0001 | NS | 0.49  (68) |
| 65-74 | Insufficient data for regression^c^ | | | | | N/A |

^c^Less than 10 States had available overdose mortality data in both males and females (see Methods and eTable 2).

^d^State level of past year psychostimulant misuse was from NSDUH, item “AMMEPDPYMU” (2018-2019), log rate / 100,000, per sex.

| **eTable 5.4**  **Cocaine** | **Significant β (beta) parameter Estimates [95%CI] and p-value** | | | | | Model R^2^ (DF) |
| --- | --- | --- | --- | --- | --- | --- |
| Age bin | Sex (Men);  Women as reference | State Median  Household  Net Worth | State  %White | State  %Black | State and sex  past year stimulant misuse  (log rate)^e^ |  |
| 15-24 | β**=**0.22  [0.098-0.34]  p=0.0008  eta^2^=0.17 | NS | NS | NS | β**=**0.49  [0.11-0.87]  p=0.013 | 0.63  (30) |
| 25-34 | β**=**0.24  [0.061-0.41]  p=0.0091  eta^2^=0.041 | β**=**1.32 X 10^-6^  [2.28 X 10^-7^ –  2.42 X 10^-6^]  p=0.019 | β**=**0.015  [0.0046-0.025]  p=0.0048 | β**=**0.015  [0.0036-0.027]  P=0.011 | NS | 0.37  (61) |
| 35-44 | β**=**0.24  [0.040-0.44]  p=0.019  eta^2^=0.074 | NS | β**=**0.020  [0.0086-0.031]  p=0.0008 | β**=**0.024  [0.011-0.036]  p=0.0005 | NS | 0.32  (62) |
| 45-54 | β**=**0.31  [0.12-0.49]  p=0.0018  eta^2^=0.10 | β**=**1.62 X 10^-6^  [3.67 X 10^-7^ –  2.88 X 10^-6^]  p=0.012 | β**=**0.017  [0.0062-0.028]  p=0.0026 | β**=**0.023  [0.012-0.035]  p=0.0002 | NS | 0.42  (61) |
| 55-64 | β**=**0.31  [0.13-0.50]  p=0.0012  eta^2^=0.11 | β**=**2.01 X 10^-6^  [7.92 X 10^-7^ –  3.22 X 10^-6^]  p=0.0017 | β**=**0.015  [0.0042-0.025]  p=0.0068 | β**=**0.024  [0.013-0.034]  p<0.0001 | NS | 0.51  (54) |
| 65-74 | Insufficient data for regression^c^ | | | | | N/A |

^c^Less than 10 States had available overdose mortality data in both males and females (see Methods and eTable 2).

^e^State level of past year cocaine misuse was from NSDUH, item “COCYR” (2018-2019), log rate / 100,000 per sex.

**eFigure 1:** Overdose mortality due to synthetic opioids *and* psychostimulants in the same person: Upper panel: all ages combined; each symbol (open circles) is one jurisdiction. Spearman correlation for men and women in each jurisdiction is included. The red line indicates the best-fit simple linear regression. Lower panel: Overdose mortality in males and females, due to synthetic opioids *and* psychostimulants, for consecutive 10-year age bins. Log-transformed data were analyzed with a univariate mixed-effects ANOVAs (sex X age bin). The label “excl.” shows that the age bin was excluded from analysis due to insufficient available data. Main effects of sex (F[1,113]=934.6), age bin (F[4,160]=19.37 and a sex x age bin interaction (F[4,113]=10.16) were found ( all p<0.0001). Bonferroni post-hoc tests are shown for sex x age bin (**** is p<0.0001).

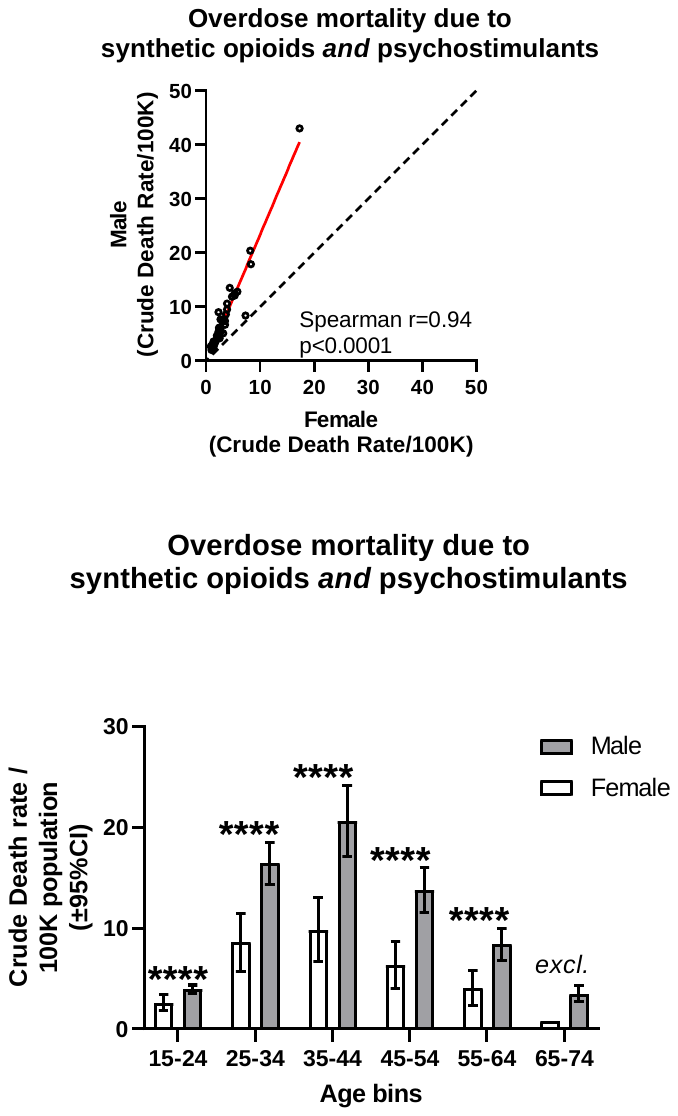

**eTable 6: Overdose mortality rates for synthetic opioids *and* psychostimulants in the same person**

**(total 51 jurisdictions)**

| **Age**  **(years)** |  | **Synthetic opioids and**  **Psychostimulant** | |
| --- | --- | --- | --- |
|  |  | **Male** | **Female** |
| **All ages**  **(range: 15-74)** | **Mean^a^ (±95%CI)** | 7.2  (5.3-9.2) | 3.3  (2.5-4.2) |
|  | **N^b^** | 47 | 43 |
|  | **Missing^c^** | 4 | 8 |
| **15-24** | **Mean^a^ (±95%CI)** | 3.9  (3.0-4.8) | 2.6  (1.8-3.4) |
|  | **N^b^** | 24 | 15 |
|  | **Missing^c^** | 27 | 36 |
| **25-34** | **Mean^a^ (±95%CI)** | 16.5  (12.3-20.7) | 8.6  (5.7-11.5) |
|  | **N** | 41 | 30 |
|  | **Missing^b^** | 10 | 21 |
| **35-44** | **Mean^a^ (±95%CI)** | 20.6  (13.6-27.7) | 9.9  (6.8-13.1) |
|  | **N** | 39 | 31 |
|  | **Missing^b^** | 12 | 20 |
| **45-54** | **Mean^a^ (±95%CI)** | 13.8  (9.3-18.3) | 6.4  (4.1-8.6) |
|  | **N** | 34 | 27 |
|  | **Missing^b^** | 17 | 24 |
| **55-64** | **Mean^a^ (±95%CI)** | 8.4  (5.2-11.6) | 4.1  (2.3-5.9) |
|  | **N** | 27 | 15 |
|  | **Missing^b^** | 24 | 36 |
| **65-74** | **Mean^a^ (±95%CI)** | 3.5  (95%CI not calculated) | 0.8  (no 95%CI) |
|  | **N** | 2 | 1 |
|  | **Missing^b^** | 49 | 50 |

^a^Overdose mortality rate per 100,000 population, unadjusted

^b^Note: If N<10 for either males or females for an age bin in a drug category, the entire age bin was excluded from analyses (both ANOVA or multiple linear regressions) for that drug category.

^c^Missing values for an age bin occurred when data for a jurisdiction were “suppressed” or considered “unreliable” in CDC WONDER

**eTable 7:** Multiple linear regressions of state-level mortality rate for overdoses caused by synthetic opioids *and* psychostimulants

|  | **Significant** β **Parameter Estimates [95%CI] and p-value** | | | | | | Model R^2^ (DF) |
| --- | --- | --- | --- | --- | --- | --- | --- |
| Age bin | Sex (Men);  Women as reference | State Median  Household  Net Worth | State  %White | State  %Black | State and sex  past year opioid misuse (log rate)^a^ | State and sex  past year stimulant misuse (log rate)^b^ |  |
| *All ages: 15-74* | *β=0.37*  *[0.24-0.50]*  *p<0.0001*  *eta^2^=0.23* | *β=****-****1.43 X 10^-6^*  *[-5.28 X 10^-7^ –*  *-2.34 X 10^-6^]*  *p=0.0023* | *NS* | *NS* | *NS* | *NS* | *0.42*  *(81)* |
| 15-24 | NS | NS | NS | NS | NS | NS | 0.31  (31) |
| 25-34 | β=0.34  [0.21-0.48]  p=<0.0001  eta^2^=0.23 | β=**-**1.21 X 10^-6^  [-1.45 X 10^-7^ –  -2.28 X 10^-6^]  p=0.027 | β=0.016  [0.0059-0.026]  p=0.0023 | NS | NS | NS | 0.50  (54) |
| 35-44 | β=0.35  [0.19-0.52]  p<0.0001  eta^2^=0.18 | β=**-**1.22 X 10^-6^  [-9.16 X 10^-9^ –  -2.44 X 10^-6^]  p=0.048 | β=0.021  [0.0092-0.032]  p=0.0006 | NS | NS | NS | 0.49  (53) |
| 45-54 | β=0.35  [0.18-0.51]  p<0.0001  eta^2^=0.20 | NS | β=0.015  [0.0030-0.027]  p=0.015 | NS | NS | NS | 0.45  (50) |
| 55-64 | β=0.30  [0.042-0.55]  p=0.024  eta^2^=0.12 | NS | NS | NS |  | NS | 0.26  (34) |
| 65-74 | Insufficient data for regression^c^ | | | | | | N/A |

Outcome is log-transformed overdose mortality rate / 100,000

^a^State level of past year opioid misuse was from NSDUH, item “OPINMYR” (2018-2019), log rate / 100,000 per sex.

^b^State level of past year psychostimulant misuse was from NSDUH, item “APPEPDPYMU” (2018-2019), log rate / 1000,000 per sex.

^c^Less than 10 states had available overdose mortality data in both males and females (see Methods and eTable 2).
